# Rethinking Paediatric Sepsis Care through Local Provider Voices and Lived Systems: A Mixed-Methods Study in Two Hospitals in Ghana

**DOI:** 10.1101/2025.07.15.25331569

**Authors:** Charles Martyn-Dickens, Sheila Agyeiwaa Owusu, Allysa Warling, Michelle Munyikwa, Gustav Nettey, Amundam Mancho, Maraisha Philogene, Evans Otieku, Ernestina Gambrah, John Adabie Appiah, Ahmet Uluer, Rebecca Elaine Cagnina, Emma Otchere, Maame Fremah Kotoh-Mortty, Eugene Martey, Leah Ratner

## Abstract

**Background:** Paediatric sepsis remains a significant cause of mortality in low- and middle-income countries (LMICs), where health systems are often resource-constrained. Global sepsis protocols, though effective in high-income settings, may not be well-suited to LMIC contexts.

**Methods:** We conducted a mixed-methods study in two hospitals in the Ashanti Region of Ghana: Komfo Anokye Teaching Hospital (KATH) and Presbyterian Hospital, Agogo (PreHA). Specifically, we conducted a retrospective chart review, followed by key informant interviews with clinical staff, and integrated our findings with a previously published situational analysis. Qualitative data analysis employed the Three Delays Framework and the Donabedian Model to identify locations and causes of delays in care.

**Results:** Seventy-one charts met the inclusion criteria for review, having a history of fever or hypothermia and complete vital signs documented (16 from PreHA, 55 from KATH). Despite KATH managing more severely ill patients with higher sepsis scores and longer stays, mortality rates were similar at both sites. The chart review highlighted gaps in documentation and inconsistent care processes. Key informant interviews revealed themes such as provider altruism, community financial support, and the positive role of research collaborations, while also illustrating systemic delays linked to financial and resource constraints.

**Conclusion:** Paediatric sepsis care in Ghana is influenced by complex and interconnected structural, cultural, and procedural factors. Our findings indicate that contextually adapted care pathways are crucial for improving sepsis outcomes in resource-constrained settings. Co-designed interventions, rather than wholly imported protocols, may offer a more sustainable approach to strengthening health systems in LMICs.

**What is known on this topic:**

- Paediatric sepsis is a major cause of morbidity and mortality globally, especially in LMICs.
- Standardised sepsis bundles have improved outcomes in high-income countries; however, they pose challenges during implementation in resource-limited settings.

**What this study adds:**

- Triangulated data show systemic and structural barriers drive delays in care-seeking, access, and treatment.
- Health system limitations have a more significant impact on clinical outcomes than individual hospital capacity.
- Locally rooted education, infrastructure, and participatory approaches are needed to improve sepsis care.

**How this study may affect research, policy or practice:**

- Encourages region-specific adaptations of sepsis guidelines.
- Highlights the role of research funding and community resilience in supporting resource-limited systems.
- Reinforces the need for implementation models grounded in the lived experiences of providers and patients.

## Introduction

Sepsis remains a leading cause of paediatric mortality globally, disproportionately impacting low- and middle-income countries (LMICs)^1^. Despite international efforts, such as the Surviving Sepsis Campaign^1^ and various risk stratification tools, including the quick Sequential Organ Failure Assessment (qSOFA)^2^ and the Phoenix Sepsis Score^3^, most protocols are based on evidence generated in high-income countries^4^. These tools often assume the presence of advanced diagnostics, intensive care infrastructure, and highly trained personnel, making their direct application in LMIC settings challenging.

Emerging evidence shows that efforts to address paediatric sepsis in LMICs have often focused on provider education and supporting clinical decision-making^5,6^. However, such interventions may be insufficient when implemented in isolation. Structural issues, including financial barriers, referral gaps, resource limitations, and sociocultural health beliefs, contribute to significant delays across the care continuum.^7^ As a result, children often present late in the disease process^8^, and care delivery is constrained by fragmented health systems^9^.

To understand these overlapping barriers, we propose a unified conceptual model that combines the Three Delays Framework with the Donabedian Model of healthcare quality. The Three Delays Framework, originally developed to understand maternal mortality, identifies critical points where care may be delayed: (1) deciding to seek care, (2) reaching appropriate care, and (3) receiving quality care^10^. The Donabedian Model^11,12^ provides a complementary structure to categorise these delays as issues of structure (e.g., resources, infrastructure), process (e.g., provider behaviour, workflows), and outcomes (e.g., morbidity and mortality). Together, these frameworks enable a more nuanced, systems-level understanding of how sociocultural and structural barriers interact across the entire care pathway.

This explanatory mixed-methods study explores how these barriers manifest in the care of paediatric sepsis at two hospitals in Ghana: a tertiary referral centre and a district-level hospital. By triangulating data from retrospective chart reviews, key informant interviews, and a previously published situational analysis^13^, we seek to move beyond algorithmic, imported models of care. Instead, we present an integrated, locally informed framework that emphasises the complex interaction of contextual factors in paediatric sepsis care and argues that provider education alone is insufficient to improve outcomes without addressing deeper structural and sociocultural determinants.

## Study Sites

This study was conducted at the paediatric departments of two hospitals in Ghana: Komfo Anokye Teaching Hospital (KATH) and Presbyterian Hospital, Agogo (PreHA).

## Ethical Approval

Ethical approval for this study was obtained from the Institutional Review Board of the Komfo Anokye Teaching Hospital (KATHIRB/AP/203/23). As the chart review was retrospective, informed consent from patients was waived. Key informants gave verbal consent for the qualitative interviews.

### Study components

#### Education

Modules were developed in collaboration with OPENPediatrics, peer-reviewed, and co-designed by members of the study team^14^. The training was delivered in real time with interactive sessions as well as pre- and post-tests to assess knowledge gains among frontline health staff (February 20 and March 26, 2024, at PreHA, and on October 10 and October 17, 2024, at KATH). While not all key informants participated in the educational component, the sessions contributed to broader clinical awareness. They were integrated into routine clinical operations through engagement with the leadership of both paediatric departments.

#### Situational Analysis

This component has been previously published^13^. Briefly, the situational analysis used a structured environmental scan to assess resource availability, staffing, infrastructure, and system-level readiness for paediatric sepsis care at KATH and PreHA. It employed the Donabedian model and the Consolidated Framework for Implementation Research (CFIR)^15^ to evaluate inner and outer setting determinants. These findings informed the adaptation of the educational intervention’s implementation, as well as the interpretation of both quantitative and qualitative study components.

#### Retrospective Chart Review

The chart review was conducted at both PreHA and KATH for patients presenting to the paediatric departments between January and April 2023. The inclusion criteria included paediatric patients presenting within the period, aged 3 months to 14 years, who presented with a documented fever (greater than or equal to 38 degrees Celsius) or hypothermia (less than 36 degrees Celsius). They also must have complete initial vital signs documentation. Charts were excluded if they lacked key data such as temperature, heart rate, respiratory rate, or mental status assessment. In total, seventy-one (71) charts were included for analysis (16 from PreHA and 55 from KATH).

#### Key Informant Interviews

Twelve (12) key informant interviews with frontline healthcare staff were conducted between September 2024 and May 2025 across both sites following informed consent. Five (5) key informants at KATH included Medical Officers (2), Resident Physician (1), Specialist Paediatric Nurse (1), and Senior General Nursing Officer (1). The seven (7) interviews at PreHA involved Staff Nurses (3), Senior Staff Nurses (2), Physician Assistant (1), and Staff Midwife (1) seconded to the department of child health. Participants were recruited through purposive and theoretical sampling to capture a diverse range of clinical roles, perspectives, and experiences. Both paediatricians at PreHA were part of the research team and refrained from participating; therefore, those recruited at PreHA represented mid-level frontline staff. The inclusion criteria were employment in clinical teams at either study site and direct experience caring for paediatric patients admitted with sepsis. Interviews were conducted either person or virtually, using a semi-structured guide, lasting about thirty (30) minutes. A token amount of 50 Ghana Cedis (3.5 USD) was provided to compensate KIIs for their time. The interviews were conducted in English. Transcripts were subsequently coded using a hybrid approach that combined deductive and inductive thematic analysis.

### Study Design Evolution

This study was initially designed as a Type I hybrid effectiveness-implementation trial^16^ to evaluate the impact of a paediatric sepsis education intervention on provider knowledge and clinical outcomes. The situational analysis and educational intervention were conducted first and informed subsequent study components. Findings from the situational analysis highlighted significant prehospital factors affecting care pathways. In response, the research team assessed the chart review data. This assessment revealed substantial missing data, limiting the ability to evaluate outcomes quantitatively. As a result, the key informant interviews (KIIs) were expanded to serve as a more central component of the study. Additional discussions confirmed that assessing intervention effectiveness alone would not fully address the research focus. Pre- and post-tests were conducted but are not reported in this manuscript. Not all participants in the education sessions were included in the qualitative interviews. Following the mid-point review, the study was restructured as an explanatory mixed-methods design utilising a Good Reporting of a Mixed-Methods Study (GRAMMS) framework.

### Conceptual Framework

To better understand how barriers to sepsis care manifest and interact in resource-constrained environments, we applied an integrated framework^17^ which draws upon the Three Delays Framework and a health systems dynamics perspective to map paediatric illness trajectories across the care pathway, from symptom onset to facility-based treatment, while analyzing how both individual/community decisions and health system functionality shape each phase.

We sought to understand how systemic and contextual elements shape clinical care. We adapted the Three Delays framework, commonly used to evaluate access to maternal and emergency care, to represent the sequential barriers patients face: (1) the decision to seek care, (2) the ability to reach appropriate care, and (3) the quality and timeliness of care once at a facility. To avoid analyzing these delays as isolated events, we embedded this pathway within a health systems lens informed by the Donabedian model of quality care: structure (e.g., resources, staffing, infrastructure), process (e.g., documentation, diagnostics, clinical protocols), and outcome (e.g., mortality, recovery, delays in care).

This integrated approach enabled us to identify not only *when* and *where* delays occur, but *why* they persist, understanding challenges in sepsis care as emerging from the interaction of cultural norms, informal coping strategies, infrastructure limitations, provider workloads, and financing models. Rather than treating patient-level decisions and facility-based care quality as distinct domains, we conceptualised sepsis care as a continuum in which barriers are cumulative, nonlinear, and systemically embedded.

The framework served as a guide for both data collection and analysis, organising our triangulated findings, i.e., retrospective chart review, key informant interviews, and a previously published situational analysis, into a cohesive systems narrative. A visual representation of this integrated care pathway is shown in Figure 2.

**Figure 1:**
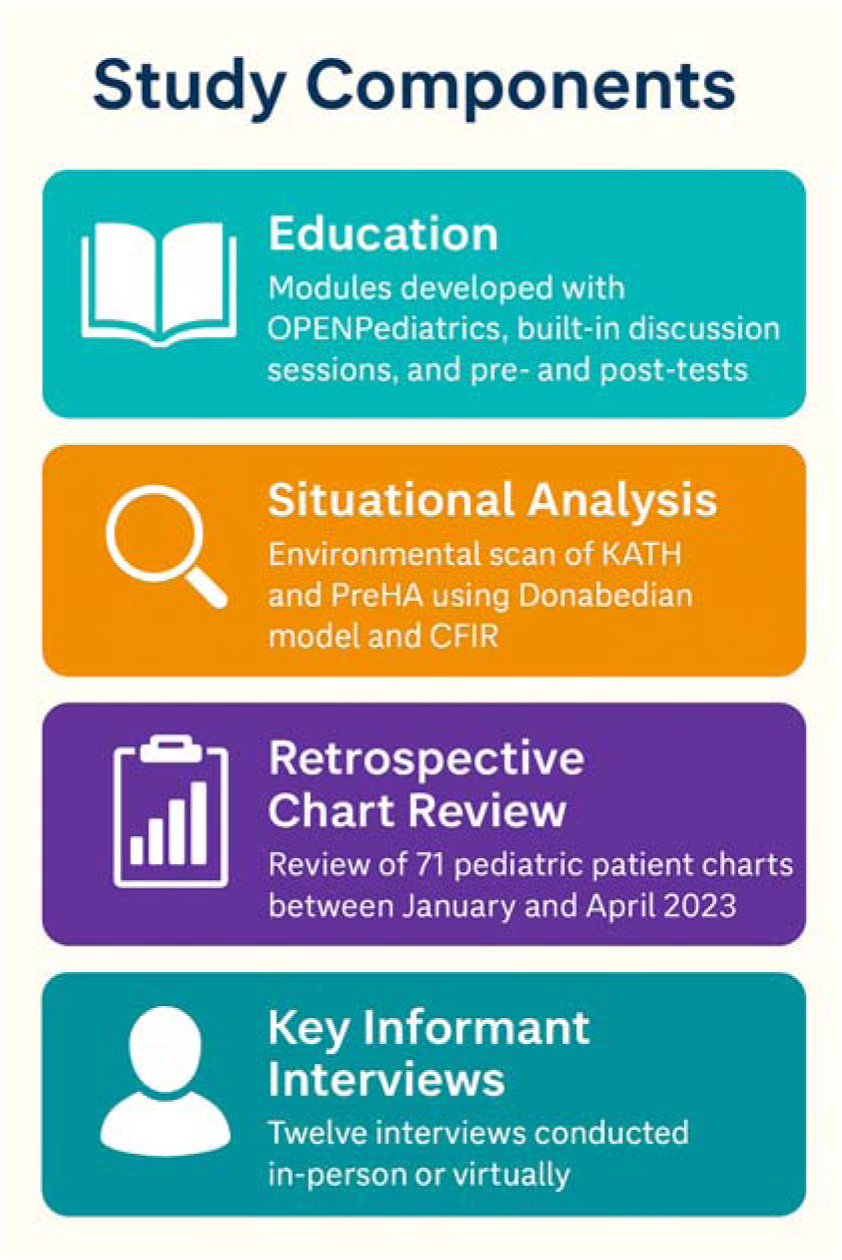
Study Components

**Figure 2:**
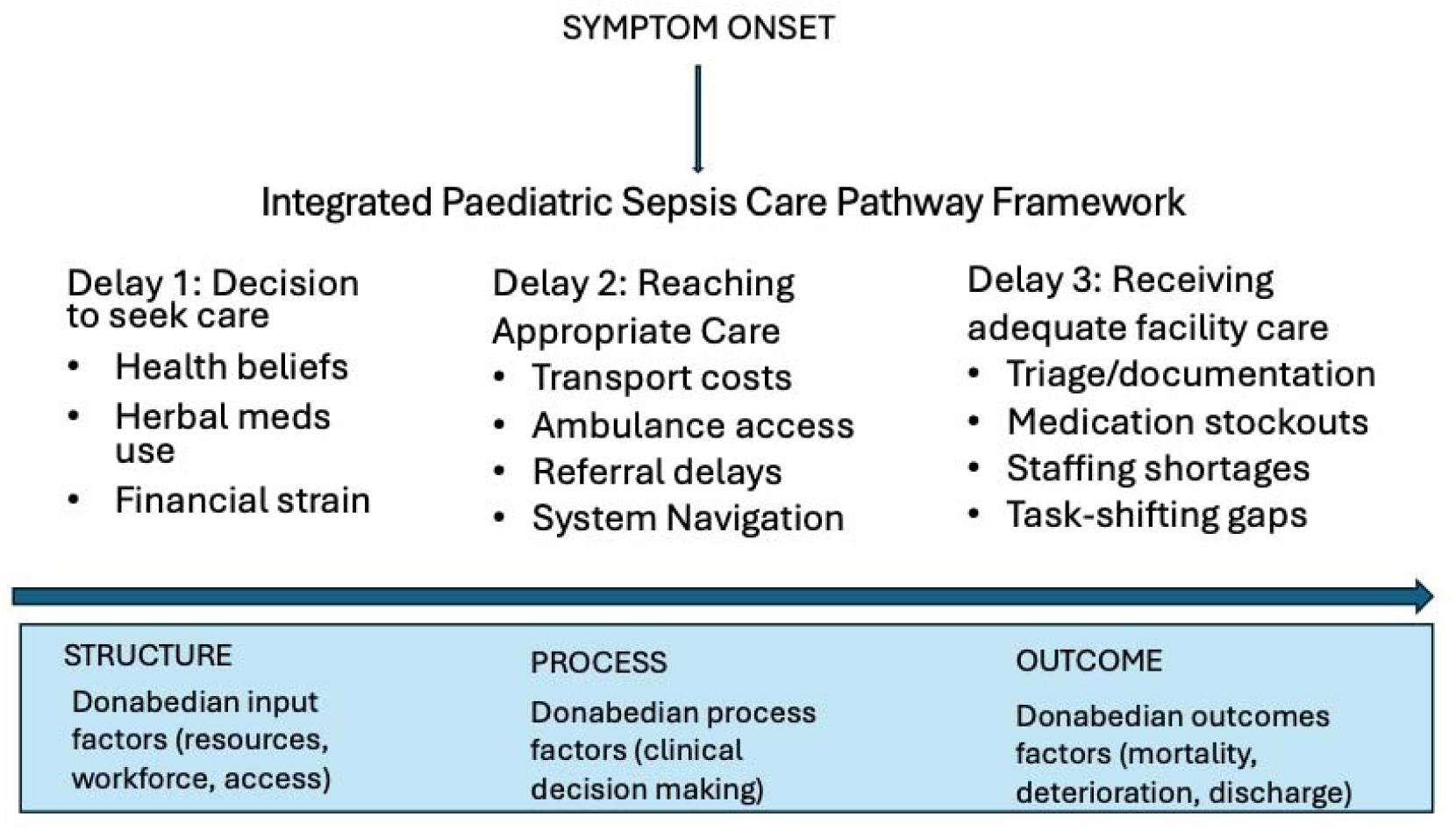
Integrated Framework

## Methods and Collection

Quantitative data were obtained through retrospective chart reviews conducted at two health facilities, and the data was disaggregated by site. This provided descriptive insights into patient demographics, sepsis severity (as measured by the qSOFA score), and clinical outcomes. The qualitative component added critical context through triangulation, helping to interpret patterns and limitations in the chart data.

qSOFA was selected a priori as a pragmatic severity score, given its reliance on bedside clinical parameters and the availability of vital signs in most records. While qSOFA has limited sensitivity in paediatric populations^2^, it was feasible for use in this setting. During the study period, the Phoenix Sepsis Score was published^3^, and we attempted to apply it retrospectively to enhance risk stratification.

However, core components of the Phoenix score, such as lactate, D-dimer, and mechanical ventilation status, were frequently undocumented. Due to these data limitations and infrastructure constraints, the Phoenix Sepsis Score was deemed unsuitable for this context.

Given the frequent lack of paediatric blood pressure monitoring due to a shortage of appropriate cuff sizes, we modified the qSOFA score by substituting hypotension with tachycardia, a well-established early marker of paediatric clinical deterioration. This modification is consistent with the Paediatric Sepsis Consensus Conference and Surviving Sepsis Campaign guidelines, which recommend adapting diagnostic tools for paediatric populations and local contexts^1,4^. Fever was also added based on clinical input from local providers, as it was commonly used as a marker of sepsis in practice. The resulting modified qSOFA included four components: fever, tachycardia, tachypnea, and altered consciousness (as assessed by Glasgow Coma Scale). Each was scored as one (1) point, resulting in a total score ranging from 0 to 4.

Scores were based on initial assessments, with a repeat measurement taken 24 hours after presentation. The highest score was used in the analysis.

Initially, the study aimed to collect a wide range of variables, based on input from a multidisciplinary team. These included: ethnicity, distance from hospital, age, sex, mid-upper arm circumference (MUAC), nutritional status, duration of admission, duration of illness prior to admission, presumed initial diagnosis, referral status, interventions prior to and within the first hour of admission, provider types, initial and follow-up sepsis scores, early diagnostics, timing of lab result return, and hospitalization outcomes.

However, due to inconsistent documentation, many variables could not be reliably extracted. After a mid-point review and further discussion with the team, the analysis was limited to charts with sufficiently complete data. These included: age, sex, sepsis severity score (highest in the first 24 hours), length of stay, duration of illness before presentation, duration of admission, and outcome (death or discharge).

Qualitative data were collected through recorded semi-structured interviews, which were conducted in English and subsequently transcribed. These key informant interviews explored provider knowledge, workflows, and system-level barriers to care. Interviews were guided by a semi-structured tool that focused on the provider’s role, clinical experiences with paediatric sepsis, and perceived enablers and obstacles to high-quality care. Participants were asked to describe both successful and challenging cases, as well as gaps in personnel, infrastructure, or system processes. They were also encouraged to identify strengths in current sepsis care practices. These insights were essential for understanding real-world challenges and informing future improvements, especially in low-resource settings.

## Analysis

### Quantitative Analysis

All statistical analyses were performed using STATA version 16. Continuous data (age, sepsis severity score, length of stay, and duration of illness) were stratified by hospital, assessed for normality, and summarised as means with 95% confidence intervals (CI). The mean difference between groups was estimated using an independent sample t-test with a two□sided significance level of 5%. Categorical variables (sex and hospitalisation outcome) were presented as counts and percentages.

The pooled quantitative results indicate that patients who died before discharge had a comparatively higher mean sepsis score than those discharged alive. The estimated mean difference of 0.9 [95% CI: 0.5 – 1.3] was statistically significant. Although discharged patients had a relatively longer length of stay (LOS) than those who died, the estimated mean difference in LOS was not statistically significant. The proportion of male discharged patients was higher than that of female patients.

#### Qualitative Analysis

The qualitative data were analysed using a thematic analysis approach, guided by an integrated framework mapping paediatric sepsis care pathways across three domains, i.e. care-seeking, access, and treatment (Three Delays Framework), while examining how each was shaped by contextual (social, cultural, logistical) and structural (infrastructure, personnel, financing) factors. The ‘structure-process-outcome’ lens of the Donabedian model informed the analysis of care delivery within each delay.

Two researchers (LR and CMD) independently reviewed the transcripts to develop an initial codebook, which was refined through discussion and consensus. Transcripts were then coded using Taguette.

Emergent themes and sub-themes were identified and organised to capture patterns and insights relevant to the research questions. Discrepancies were resolved through team discussion until consensus was reached. Thematic saturation was considered achieved when no new themes emerged from the data, which was reached after four (4) interviews.

#### Triangulation

To enhance the rigour and credibility of the findings, we applied methodological triangulation by integrating quantitative chart review data with qualitative interviews through the lens of our previously published situational analysis. The quantitative data (including clinical characteristics and outcomes) were examined alongside qualitative themes (such as perceptions of delays, clinical care quality, and systemic barriers) to identify areas of convergence, divergence, and complementarity. Triangulation was conducted during the analysis phase by comparing quantitative results (e.g., differences in sepsis scores, length of stay) with qualitative narratives (e.g., descriptions of delays and clinical decision-making). This approach enabled a more nuanced understanding of patient outcomes and the contextual factors that influence them. Discrepancies were discussed within the multidisciplinary research team until consensus was achieved, ensuring a more comprehensive and credible interpretation of the data.

#### Results

A total of 931 paediatric admissions occurred across both hospitals between January and April 2023 (121 in PreHA; 810 in KATH). Charts from 71 children were included in the review (16 from PreHA, 55 from KATH) [Table 1]. Data on reasons for exclusion were not collated. The mean duration of illness prior to presentation was 12 days. The mortality rate was similar across both sites (~32%), despite patients in KATH having higher average sepsis scores and longer hospital stays. The majority of participants were male (61%) with an average age of 43 months at PreHA and 39 months at KATH. Patients at KATH had longer hospital stays (mean: 6.8 days) than those at PreHA (mean: 3.9 days), and higher average sepsis severity scores (2.9 vs. 2.4).

**Table 1a:** Chart review findings.

|  | **PreHA**<br>**Mean (95%CI)** | **KATH**<br>**Mean (95%CI)** | **Overall**<br>**Mean (95%CI)** |
| --- | --- | --- | --- |
| Age in months | 43 (20 – 65) | 39 (28 – 51) | 40 (30 – 50) |
| Sepsis severity score (qSOFA) | 2.4 (1.9 – 2.9) | 2.9 (2.7 – 3.1) | 2.8 (2.6 – 3.0) |
| Length of stay (in days) | 3.9 (1.4 – 6.5) | 6.8 (5.1 – 8.4) | 6 (5 – 8) |
| Duration of illness prior to presentation (in days) | 12 (-0.15 – 23.9) | 13 (4 – 21) | 12 (5 – 20) |
| Sex | N (%) | N (%) | N (%) |
| Female | 8 (50.0) | 20 (36.4) | 28 (39.4) |
| Male | 8 (50.0) | 35 (63.6) | 43 (60.6) |
| Total | 16 (100.0) | 55 (100.0) | 71 (100.0) |
| Outcome | N (%) | N (%) | N (%) |
| Death | 6 (37.5) | 17 (30.9) | 23 (32.4) |
| Discharge | 10 (62.5) | 38 (69.1) | 48 (67.6) |
| Total | 16 (100.0) | 55 (100.0) | 71 (100.0) |

**Table 1b:**
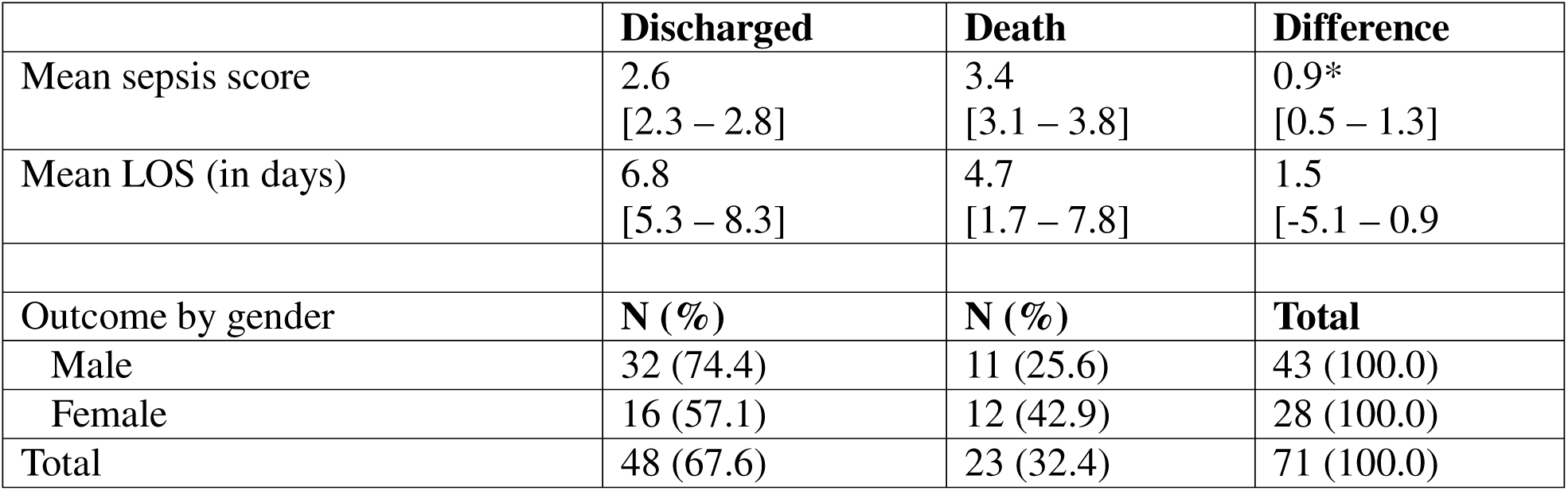
Disaggregated outcome by gender, sepsis score, and length of stay.

Key informant interviews (n=12) revealed systemic challenges including a reliance on herbal remedies, financial constraints, transportation barriers, fractured referral systems, supply shortages, and documentation gaps. Emergent themes included the role of community support, physician altruism, and the impact of research partnerships on care delivery.

**Table 2:** Qualitative Results.

| **Parent Theme** | **Sub Theme** | **Quote (with citation)** | **Relation to the Model** |
| --- | --- | --- | --- |
| **Delay 1: Delay in deciding to seek care** | Herbal medication use | PreHA interview 3:<br>*“In fact, those herbs, those herbs around them, they mostly use them. They think it will help. By the time they get here, the situation is very worse.”* | Three Delays Model: Cultural beliefs and a reliance on traditional medicine contribute to delays in recognizing severity and seeking care.<br>Donabedian: Structural limitation (lack of trust or access to formal care). |
| **Delay 1: Delay in deciding to seek care** | Pharmacies as a first stop | PreHA Interview 3:<br>*“Sometimes they... You attend to the pharmacy, the drug stores. All that they know, oh, my child is sick, it's malaria. So they just go and buy this malaria [drugs]. Maybe even it's anemia, they don't know. Something serious has happened, they don't know. All that they know is malaria. They just go to the nearby clinic and go and buy malaria medication.”* | Three Delays Model: Misdiagnosis and inappropriate early care delay escalation.<br>Donabedian: Process failure in informal care, structural lack of accessible diagnostic services. |
| **Delay 1: Delay in deciding to seek care** | Financial strain | KATH Interview 4:<br>*“They mostly do over-the-counter medications, possibly have all medications, and when they realize that now it's overwhelming, then they can report to a healthcare facility”* | Three Delays Model: Financial barrier delays initial engagement with the formal health system.<br>Donabedian: Structural constraint related to affordability. |
| **Delay 2: Delay in reaching the health care facility** | Financial strain | PreHA Interview 4:<br>*“So sometimes, when they are in the villages and those far-far villages, what I know is that they might seek first intervention from the pharmacist and the drug store. So that will give them some minor, minor drugs to take. Like that (unclear drug name). So, when they take it and it's not going, that's whereby they might come. Or, as I was first saying, if I come to there, maybe they make me buy ceftriaxone and tomorrow they change ceftriaxone. And see that it's waste of money.”* | Three Delays Model: Economic burden deters consistent and escalated care-seeking.<br>Donabedian: Process issues (changing prescriptions); Structure (cost). |
| **Delay 2: Delay in reaching the health care facility** | Health centers giving inaccurate care | KATH Interview 1:<br>*“Yes. So, because KATH is a tertiary hospital, so sometimes, instead of the district hospital..... okay, so usually they refer cases from the district hospital to you. You may have a patient spending a lot of time or a longer time in the district hospital. With them doing the cocktail of things or not, maybe not picking up, the*<br>*sepsis. And by the time they refer that case to us, it's sort of too late.”* | Three Delays Model - Delay 2: Referral delays due to under-recognition.<br>Donabedian: Process failure in district hospitals, Outcome deterioration from late recognition. |
| **Delay 3: Delay in adequate care at the appropriate facility** | Burden of Sepsis | PreHA interview 2:<br>*“Yeah, sometimes they will take the first two doses and then the third one, just miss it. Then you come and start with the fourth day. Yeah, so there's been breakages”*<br>KATH Interview 2:<br>*“Ok. So, we all know we have specific parameters for I.O. (intraosseous access), so what we can do is improvise. But you know in our setting, lack of logistics and resources, we have to improvise to care for the patient. If you*<br>*don't have the needles for the IO, you have to use the 50cc needles. But it's not always the IO, sometimes we do the scalp or the jugular. But this is improvising for the IO."* | Three Delays Model:<br>A lack of resources due to burden (equipment, laboratory services, trained personnel, for example)<br>Donabedian: Process failures occur because of a lack of resources |

### Delay 1: Deciding to Seek Care – Caregiver Perceptions and Financial Barriers

*“They usually start with herbal medications and only come to the hospital when things have gotten really bad.”* – KATH Interview 1

Providers noted that caregivers frequently delayed seeking biomedical care for children due to cultural norms, financial difficulties, and reliance on herbal medicine. Key informants explained that herbal remedies were often the initial approach, commonly continued until a child’s condition deteriorated considerably. Chart review data corroborate this, showing an average duration of illness before presentation of 12 days at PreHA and 13 days at KATH.

Providers hypothesised that these delays were shaped by both household-level constraints (e.g., transport costs, informal user fees) and broader health system limitations (e.g., lack of accessible or affordable primary care options). Some families faced difficult trade-offs between seeking care and maintaining their income, while others delayed care due to distrust or past poor experiences with the formal health system.

### Delay 2: Reaching Appropriate Care – Transport, Referral, and Navigation Challenges

*“By the time they go to the health centre and then are referred to us, the child is often already in shock.”* – PreHA Interview 2

Even when the decision to seek care was made, systemic gaps in transportation and referral networks impeded timely access to appropriate facilities. Participants described ambulance shortages, high transport costs, and long travel times, particularly in rural areas. These barriers were compounded by delays at lower-level health centres, where initial recognition of sepsis was limited, and referral processes were slow or unclear.

This resulted in prolonged illness trajectories and, in some cases, further deterioration en route. The absence of integrated referral systems led to care fragmentation and an increased caregiver burden.

### Delay 3: Receiving Adequate Facility-Based Care – Resource and Process Constraints

“*Sometimes they will run away and not pay for the labs, or we can’t find the forms to request them, and we just have to guess*.” – PreHA Interview 1

At both study sites, once patients arrived at the hospital, care was influenced by a combination of structural and procedural limitations. Key informant interviews revealed that providers possessed a strong understanding of sepsis and its treatment. They identified early recognition, prompt antibiotic administration, and hemodynamic support as critical strategies. However, chart reviews showed inconsistent documentation of vital signs, sepsis indicators, and treatment timelines, as well as irregular use of severity scoring tools. This disconnect was not due to a lack of clinical knowledge but rather stemmed from structural constraints such as heavy workloads, documentation burdens, and the absence of standardised protocols. Furthermore, shortages of antibiotics, lack of paediatric-specific equipment, and high provider workloads limited the effectiveness of sepsis management protocols.

Providers described both moral distress and improvisation in navigating resource constraints. Some relied on personal or community resources to support patient care. Interviewees also noted that despite clinical knowledge often being strong, the inability to act due to missing laboratory results, delayed diagnostics, or inadequate staffing meant this knowledge could not be translated into timely care.

### Emergent Themes: System Workarounds and Provider Resilience

“*We do what we can. Sometimes our decisions are right, sometimes not—but we can’t wait for labs that won’t come*.” – KATH Interview 3

Despite these systemic challenges, providers demonstrated resilience through informal solutions, including conducting bedside assessments without laboratory tests, relying on collective clinical experience, and occasionally fundraising to support families. Notably, patients participating in research programs gain access to essential laboratory services, medications, and enhance their practice.

However, reliance on these workarounds was neither sustainable nor equitable, underscoring the need for more structural and systemic solutions to these problems.

### Results of Triangulation

**Table 3:** Framework-Based Triangulation Matrix.

| Framework Domain | Chart Review Findings | Key Informant Interview Findings | Situational Analysis | Integrated Interpretation |
| --- | --- | --- | --- | --- |
| Delay 1: Deciding to Seek Care | Mean duration of illness before presentation: 12–13 days | PreHA Interview 3<br>Caregivers try home remedies or wait due to stigma, cost, and misinformation<br>*“They mostly do over-the-counter medications... By the time they get here, the situation is very worse.”* | At KATH, patients arrive after visiting other facilities; referral delays are likely<br>PreHA patients often first access herbal medicine or pharmacies | Delay in seeking care is shaped by financial strain, when traditional medicine is seen as an alternative to Western medicine for sepsis, and systemic distrust; education and community engagement are key |
| Delay 2: Reaching the Facility | Not captured | Transport and funds are lacking; long journeys and weak referral systems noted | KATH has a more structured referral system<br>PreHA depends on family-arranged/paid-for transport to KATH (2-3 hours’ journey); no PICU | Barriers to reaching care include geographic inaccessibility (long distances to care and road challenges), constrained emergency transport, and cost burdens |
| Delay 3: Receiving Adequate Care | Incomplete documentation; delayed vital sign recording; care plans inconsistent | Delays due to supply shortages, workflow inefficiencies | KATH has a PICU, PEU, and triage systems<br>PreHA lacks a PICU; limited staff, poor coverage; frequent stock-outs | Systemic workflow obstacles due to resource constraints |
| Structure<br>(Donabedian) | Not applicable | PreHA Interview 4<br>*"We need more pulse oximeters... if more than 3 babies come..."* | KATH has 69 pediatric beds and 75 physicians in department of child health<br>PreHA has 61 pediatric beds and 2 pediatricians; research funding supports diagnostics<br>Both have frequent power outages, but reliable generators | Staffing, space, diagnostics, and supply chain gaps compromise care. Differences in structural capacity, shaped by historical underfunding and reliance on research partnerships, inform interventions. Reliance on research also allows for increased capacity and diagnostics, however. |
| Process<br>(Donabedian) | Extended length of stay (mean 6.8 days at KATH) | Triage varies; lack of standardized responses | Global guidelines are misaligned with local realities.<br>Systemic financial gatekeeping.<br>workflow bottlenecks at PEU/PICU levels | Global protocols must be adapted for local contexts, reflecting logistical and financial constraints and hospital specific workflow dynamics |
| Outcome<br>(Donabedian) | Mortality ~32%; similar across both sites despite higher acuity at KATH | Providers believe deaths would be preventable with earlier presentation and support | Sepsis remains a leading cause of pediatric death; systemic inefficiencies are noted as likely contributors | Mortality is more reflective of systemic and structural challenges than clinical capability; improving structure and process may reduce preventable deaths |
| Emergent Theme:<br>Financial Strain | Not applicable | Providers cite inability to pay as a significant barrier to care | Many diagnostics and medications require upfront payment; treatment is delayed as families raise funds | Financial barriers are deeply embedded and delay all stages of care; addressing cost structures and insurance coverage is essential |
| Emergent Theme:<br>Community Support / Physician Altruism | Not applicable | Providers and families pool resources to support children | Community resilience is present, but physician-led resource support is not sustainable without system-level solutions | Informal support networks exist but are fragile; structural investment is needed to move from ad hoc to sustainable care models |
| Emergent Theme:<br>Perceived Impact of Research Presence | Not applicable | Staff noted increased attentiveness and access during research engagement | Partnerships with PreHA (Kumasi Centre for Collaborative Research, German collaborators) improve clinical capacity; Health care support due to research funding | Research involvement can strengthen systems but may create dependency and inequity across facilities; future partnerships must promote system-wide equity |
| Emergent Theme:<br>Education Improves Outcomes | Not captured | Training increases provider confidence and patient trust | Prior OPENPediatrics modules addressed gaps; future education modules planned | Provider and caregiver education are scalable strategies that support early recognition and appropriate care, especially in resource-constrained settings |
| Emergent Theme:<br>Herbal/<br>Traditional<br>Medication Use<br>and Pharmacies as<br>the first point-of-<br>call | Not directly<br>captured | PreHA Interview 3<br>*"In fact, those*<br>*herbs... they mostly*<br>*use them. They*<br>*think it will help. By*<br>*the time they get*<br>*here, the situation*<br>*is very much*<br>*worse."* | Seen as a primary care<br>access point in rural areas,<br>such as those served by<br>PreHA | Traditional systems are<br>part of the health-seeking pathway;<br>integration and<br>education are needed to<br>promote earlier referral<br>and trust in biomedical<br>care |

## DISCUSSION

This study moves beyond traditional analyses of paediatric sepsis as a clinical or educational problem and instead frames it as a care pathway issue shaped by complex, interacting systems. By integrating chart reviews, provider narratives, and environmental analyses within a unified framework, we demonstrate that delays in paediatric sepsis care in Ghana are not discrete failures, but rather the result of systemic fragmentation and contextual realities. Our findings demonstrate that barriers at the household level (e.g., delayed decision-making due to financial constraints or reliance on herbal remedies) interact with logistical gaps (e.g., poor referral networks, ambulance shortages) and facility-level constraints (e.g., documentation gaps, medication stockouts, staff shortages) to produce compounding risk.

Our qualitative findings align with existing evidence from Ghana and the broader Sub-Saharan African region, showing that caregivers frequently seek traditional healers or pharmacy-based care as their initial response to paediatric illness, motivated by perceived affordability, cultural preference, and accessibility^18^. Studies in Ghana revealed that nearly 50% of surgical patients initially consulted herbal or religious healers^19^, often resulting in delayed, advanced presentations^20^. Similarly, a Ghana Demographic and Health Survey analysis found that caregivers often started with pharmacy or over-the-counter treatments for paediatric respiratory illnesses, delaying formal clinical care, especially when cost or accessibility issues were present^21^. These patterns reinforce the notion that economic and cultural factors significantly drive delays in sepsis care, resulting in patients presenting in more critical condition.

Financial support for hospitalised children in Ghana is primarily provided through the National Health Insurance Scheme (NHIS), which was implemented in 2003 as a step toward universal health coverage. An exemption policy exists to enable the poor and vulnerable to access healthcare^22^. Despite this social intervention, out-of-pocket payments for ambulance services, as well as some critical laboratory and pharmaceutical services, disincentivise health-seeking behaviours^23^.

Though the introduction of improved ambulance services has been associated with increased utilization of emergency transportation to health centres,^24^ its benefit is unlikely to be substantial with population density and poor road networks^25^.

Despite these challenges, broader cultural values in West African contexts, which prioritise interdependence and communal responsibility, nonetheless exist. As shown by Agulanna and colleagues, collectivistic orientations shape social behaviours around health and well-being,^26^ while the concept of African communalism, ‘I am because we are’^27^, underscores an ethic of mutual support. These cultural norms help explain why providers and community members at PreHA engaged in personal and communal financial efforts as part of sepsis care. This is reinforced by historical examples—such as during the Ebola epidemic—where West Africans “stood by the ill” even under risk.^28^ Collectively, these data suggest that provider-level altruism and community solidarity are not isolated phenomena but are deeply embedded in local social and ethical systems.

This study reframes paediatric sepsis care in Ghana not as a failure of knowledge or guideline adherence, but as the outcome of a fragmented, under-resourced system shaped by structural and sociocultural forces. By integrating the Three Delays Framework with the Donabedian model within a unified care pathway framework, we demonstrate how delays accumulate across the continuum of care, with provider capacity constrained by systemic forces that extend far beyond individual decision-making.

The findings reveal that clinical knowledge among providers was often strong; however, systemic limitations prevented its consistent translation into timely care. Delays in seeking care were deeply rooted in economic vulnerability, reliance on traditional healing systems, and a lack of trust in biomedical interventions. Transportation failures and inadequate safety nets created barriers to accessing care, while infrastructure gaps, stockouts, and referral bottlenecks hindered care delivery once patients arrived at facilities.

These insights challenge the prevailing assumption that provider education alone can improve paediatric sepsis outcomes in low- and middle-income countries. While knowledge was frequently present, the ability to act upon it was repeatedly undermined by cost barriers, resource constraints, and system fragmentation. Sustainable improvements in paediatric sepsis outcomes will therefore require regionally adapted, systems-informed strategies that address both the structural foundations of care and the lived experiences of those delivering and receiving it, moving beyond educational interventions to comprehensive system strengthening.

Rather than isolated events, these barriers were cumulative and interconnected. For example, financial barriers influenced both the decision to seek care and the ability to follow through with treatment recommendations. Likewise, a delay in reaching appropriate care increased the clinical severity on arrival, placing further strain on already limited facility resources.

Yet, despite these challenges, providers demonstrated remarkable resilience and ingenuity. Their improvisation in the face of systemic constraints highlights both the need for structural investment and the risk of moral distress in contexts where the burden of compensating for system failures falls on frontline staff.

Ultimately, addressing paediatric sepsis in LMICs requires more than algorithms or educational workshops. It demands investments in the health system’s backbone: referral coordination, supply chains, financing mechanisms, and supportive policy environments.^29^ Future interventions must be locally adapted, co-designed with providers, and grounded in the lived complexity of care.

### Limitations

This study has several limitations. First, the retrospective nature of the chart review limited the completeness and consistency of data, particularly in vital sign documentation and clinical decision-making pathways. Additionally, data collectors varied over time, which may have introduced inconsistency in abstraction despite the use of standardised forms. Charts from only January to April 2023 were reviewed; therefore, there is a potential risk of seasonal bias. However, this was the same time frame across both sites, which mitigates this variation. The sample size was relatively small, particularly at PreHA, which limits the generalizability of the quantitative findings. Our focus, however, was not on the burden of sepsis but on the structures, processes and outcomes of sepsis care. Moreover, while the qualitative component captured rich insights from providers, it did not include the perspectives of patients or caregivers. Nonetheless, provider narratives frequently referenced barriers experienced before arrival at the hospital and between facilities, suggesting that future work should include community and pre-hospital perspectives to more fully understand the continuum of delays in sepsis care.

### Conclusion

This study underscores the importance of contextualising paediatric sepsis care within the realities of health systems in low-resource settings. By triangulating chart review data with provider perspectives through the lenses of the Three Delays Framework and the Donabedian Model, we identified critical barriers that span both clinical and structural domains. Our findings highlight that delays in seeking, reaching, and receiving care are not isolated events but rather interconnected challenges rooted in systemic constraints, inadequate infrastructure, and limited resource availability.

Although our initial efforts focused on evaluating clinical outcomes through a traditional effectiveness lens, the study’s transition to an explanatory, mixed-methods approach revealed the deeper complexities that influence paediatric sepsis care. These insights emphasise the importance of developing facility-specific sepsis pathways that are not only evidence-informed but also grounded in the lived experiences of both providers and patients. Ultimately, improving sepsis outcomes in Ghana and similar settings requires moving beyond the importation of externally derived protocols toward co-designed, context-responsive interventions, pre-, peri- and post-hospital, that acknowledge both clinical science and local realities.

## Data Availability

All data produced in the present work are contained in the manuscript

## Acknowledgements

We extend our heartfelt gratitude to all key informants who generously shared their time and insights. We are deeply honoured to learn from their experiences and remain committed to improving health systems and structures in their honour.

GRAMMS Checklist: Mixed-Methods Study on Paediatric Sepsis Care in Ghana

O’Cathain A, Murphy E, Nicholl J. The quality of mixed methods studies in health services research. *J Health Serv Res Policy*. 2008 Apr;13(2):92–8. doi:10.1258/jhsrp.2007.007074

1. Explain the rationale for using a mixed methods design The study originally aimed to evaluate the effectiveness of a paediatric sepsis education intervention. However, due to structural and contextual challenges (e.g., poor documentation), it evolved into an explanatory mixed-methods study. Mixed methods were necessary to capture both measurable clinical data and deeper systemic and sociocultural barriers influencing paediatric sepsis care.
2. Describe the design in terms of the purpose, priority and sequence of methods The design evolved from a Type I hybrid effectiveness-implementation trial into an explanatory mixed-methods study. The sequence was: (1) situational analysis and education intervention, (2) chart review, and (3) key informant interviews. Priority shifted to qualitative methods midway, due to quantitative data limitations. Integration was performed at the analysis phase.
3. Describe each method in terms of sampling, data collection, and analysis

- Quantitative: Retrospective chart review of 71 charts (PreHA: 16, KATH: 55) from Jan–Apr 2023; included patients aged 3 months–14 years with documented fever/hypothermia. Data were analyzed with STATA v16 using t-tests and descriptive statistics.
- Qualitative: 12 semi-structured interviews with frontline staff recruited via purposive and theoretical sampling. Interviews were transcribed and thematically analyzed using a hybrid inductive/deductive approach and Taguette. Saturation was reached after 4 interviews.
4. Describe where integration occurred, how it occurred, and who participated in the integration Integration occurred during analysis, comparing quantitative chart review findings with qualitative themes and a previously published situational analysis. Triangulation revealed areas of convergence/divergence. Discrepancies were resolved via team consensus, involving both qualitative and quantitative researchers.
5. Describe any limitations of one method associated with the presence of the other method Chart review was limited by incomplete documentation and small sample size, which shifted the emphasis to qualitative methods. Conversely, qualitative interviews could not include caregivers, limiting insight into prehospital delays. Yet provider narratives often referenced community-level experiences, partially mitigating this gap.
6. Describe any insights gained from mixing or integrating methods Integration allowed for a systems-level understanding of paediatric sepsis care in Ghana. Quantitative findings highlighted documentation and severity trends, while qualitative data revealed structural delays, provider improvisation, and systemic constraints. Mixing methods validated and contextualized the observed clinical outcomes, showing that sepsis care quality is more shaped by systemic issues than clinical knowledge alone.

